# Efficacy and safety of triple versus dual combinations of antihypertensive drugs: A systematic review and meta-analysis of double-blind randomized clinical trials

**DOI:** 10.64898/2026.07.03.26356990

**Authors:** Rupasvi Dhurjati, Rashmi Pant, Gautam Satheesh, Anshika Mittal, Anthony Rodgers, Abdul Salam

## Abstract

We evaluated the blood pressure (BP)-lowering efficacy and safety of triple vs dual therapy of antihypertensive drug (AHTD) combinations, among adults with hypertension. Seventeen randomized, double-blind trials (41 comparisons and 13,461 participants) comparing triple versus dual therapy for ≥3 weeks identified by multiple literature databases searches including PubMed, Cochrane Central Register of Controlled Trials (CENTRAL) until October 2024 were included in the meta-analysis.

Triple therapy achieved a greater reduction in systolic BP (SBP) compared with dual therapy (26.9 vs. 21.7 mmHg, mean difference 5.4 mmHg [95% CI, 4.7–6.2]). Among patients receiving dual therapy at submaximal and maximal doses, the addition of a third drug further reduced SBP by 7.5 and 3.6 mmHg, respectively. BP control was significantly better with triple therapy (60% vs. 47%, RR=1.34 [1.27–1.41]). Withdrawal due to adverse events was slightly higher in the triple therapy group (4% vs. 3%, RR=1.5 [1.2–1.8]). Triple AHTD therapy provides superior BP reduction and is well-tolerated compared to dual therapy.

## Introduction

Hypertension continues to be the leading cause of global mortality. Most patients with hypertension require treatment with two or more antihypertensive drugs (AHTDs) to achieve blood pressure (BP) control and reduce morbidity and premature mortality. However, globally, most treated patients only receive treatment with one AHTD, resulting in fewer than 1 in 4 individuals achieving the target BP of <140/90 mmHg. [1, 2] The proportion needing two or more AHTDs is even greater with lower BP targets that are now widely recommended for most or all patients. [3–5]

Most hypertension guidelines advocate for initiating single-pill combination (SPC) therapy as the primary treatment for most hypertensive patients to achieve blood pressure control below 140 mmHg SBP, marking a significant advancement in hypertension management. While some guidelines recommend escalating dual therapy to the maximal dose dual combination, others advocate for adding an additional drug to optimize the dosage. [6]

A previous meta-analysis evaluated the efficacy and safety of triple and dual combination BP-lowering drug therapy. [7] However, given the increasing emphasis on combination therapy and the publication of new trials comparing triple and dual combinations of AHTDs, an updated meta-analysis is essential, including recent trials to provide up-to-date evidence. Therefore, this systematic review and meta-analysis was conducted to compare, among adults with hypertension, the efficacy and safety of triple and dual combination therapy of AHTDs for BP lowering.

## Methods

We conducted an updated systematic review and meta-analysis, following the protocol previously registered on PROSPERO (registration number: CRD42014006977).

### Literature search

We conducted a systematic search on PubMed, Cochrane Central Register of Controlled Trials (CENTRAL) from inception to October 2024 to identify eligible trials. Appropriate Medical Subject Headings and free-text terms, including keywords, were used to develop the search strategy (full search strategy is reported in **Supplementary Tables S1 and S2**). To identify unpublished trials, drug approval packages from the United States Food and Drug Administration website were also searched. Additionally, the bibliography of relevant reviews and included trials was also reviewed.

### Eligibility criteria for including trials

Trials were eligible for inclusion if they satisfied the following criteria: 1) randomised and double-blind, 2) enrolled adults (age ≥18 years) with hypertension (SBP ≥140 mmHg and or DBP ≥90 mmHg or taking antihypertensive drugs) without any significant comorbid conditions that could alter the effects of BP-lowering drugs, 3) had at least one comparison of drugs and doses of triple versus dual combinations of oral angiotensin-converting enzyme inhibitors (ACEIs), angiotensin II receptor blockers (ARBs), beta-adrenergic blockers (BBs), calcium channel blockers (CCBs), or thiazide/thiazide-like diuretics for at least 3 weeks, and 4) reported data to evaluate BP-lowering efficacy. Studies involving optional titration to dual or triple combination therapy, such that different participants received different drugs or doses, were excluded. Some trials included more than one treatment period. For these trials, the final treatment period was used for the main analysis, while all treatment periods lasting at least three weeks were included in the analysis of the effects of up-titration

### Selection of trials and data extraction

After excluding duplicates, two reviewers independently and in duplicate reviewed the title and abstract and then the full text of each record to determine eligibility for inclusion. From each included trial, relevant data on characteristics of the trial, participants, interventions and outcomes data were extracted by two reviewers, in duplicate and independently, using a structured data collection form on Distiller-SR. [8]

### Primary and secondary outcomes

The primary outcomes were reduction in SBP (difference in change in mean SBP from baseline to end of the treatment) and withdrawal of treatment due to adverse events (WDAEs). Secondary outcomes were reduction in DBP, proportion of participants achieving BP control (SBP <140 and DBP <90 mmHg), incidence of any adverse events (AAEs), and treatment-related adverse events (TRAEs).

### Assessment of methodological quality of included trials

The risk of bias of individual studies was assessed using Cochrane’s risk of bias assessment tool, Version 1.0. [9] Where appropriate, we used the results of the risk of bias assessment for sensitivity analysis. The certainty of evidence was assessed using the Grading of Recommendations, Assessment, Development, and Evaluations (GRADE) approach, and GRADEPro software was used to generate GRADE Evidence profile tables. [10]

### Data management and analysis

Doses of AHTDs were standardized based on the usual maintenance dose and the WHO-defined daily dose **(Supplementary Table S3)**. For the trials involving forced up-titration of dose, the last treatment period was taken for analysis. The mean difference between treatment groups, along with 95% confidence intervals (CI), was used to summarise reductions in SBP and DBP. The proportion of participants achieving BP control at the end of treatment, as well as the safety outcomes (WDAEs, AAEs, TRAEs, and SAEs), were summarized as a percentage of participants with at least one event along with relative risk (RR) and its 95% CI. In case of multiple comparisons of triple and dual combinations from a trial, and if a trial group contributed more than once to a meta-analysis, the number of participants (and number of events for binary outcomes) was divided by the number of times the group contributed to a meta-analysis, thus avoiding the issue of double-counting. All meta-analyses were conducted using DerSimonian and Laird random-effects model. The heterogeneity in effects was detected by the Q test and quantified by I^2^ statistics, with an I^2^ value higher than 50% considered as ‘substantial’ heterogeneity. Meta-regression analysis was conducted to evaluate the effects of baseline BP and the class of the third drug on the primary outcome. Where appropriate, funnel plots were used to investigate publication bias. Subgroup analyses were conducted to evaluate efficacy by the type of trials and the standard dose of the third drug in triple combination. Data were analysed using comprehensive meta-analysis software (V4.0) and Stata 18. [11]

## Results

### Search results

The process of identification and selection of trials is presented in the PRISMA flowchart (**Supplementary Figure S1**). Overall, 17 trials (41 comparisons, 13,461 participants) were included.

### Characteristics of the included trials and participants

Three types of trials were identified based on participants’ treatment status at baseline (see **Supplementary Table S4)**: (1) AHTD-free baseline trials (n = 6), involving participants who were either untreated or had undergone a washout of antihypertensive drugs (AHTDs) before being randomised to receive either triple or dual combination therapy; (2) Uncontrolled-on-dual trials (n = 10), involving participants whose BP was uncontrolled on dual combination therapy and who were randomised to either continue dual therapy or switch to triple combination therapy; and (3) Triple run-in trials (n = 1), involving participants who underwent a run-in period on triple combination therapy and were then randomised to either continue triple therapy or switch to dual combination therapy.

Most trials (71%) evaluated a combination of an ARB, a CCB, and a thiazide diuretic. In all trials, the dual combination included the same components as the triple combination at identical doses, except in one trial, where the ARB used in the triple combination was valsartan, and the dual combination included losartan [12]. The mean age of participants was 58 years, and the most prevalent comorbidity at baseline was diabetes mellitus (23%). BP was measured in-clinic in a seated position in all trials, and at trough in 12 trials (70%), while the remaining trials did not report the timing of BP measurement.

### Risk of bias in the included trials

Overall, none of the 17 studies had a high risk of bias in any of the six domains assessed (**Supplementary Table S5**). Several trials inadequately reported random sequence generation, allocation concealment, and blinding. These domains were judged as having a low risk of bias, as the deficiencies were most probably due to reporting issues rather than methodological flaws, considering the trial organisation and settings. Most trials were funded by industry, and in some cases, authors declared receipt of support from the sponsor. These trials were judged as having an unclear risk of bias for the ’other bias’ domain. The funnel plot based on the difference in mean SBP reduction showed only subtle asymmetry in dispersion of studies **(Supplementary Figure S2).** Egger’s test indicated some degree of publication bias or small-study effects (β_ = 1.82; p = 0.0156). A trim-and-fill analysis identified 15 potentially missing comparisons and produced an adjusted pooled estimate that was similar to the original (mean SBP difference: -5.5 vs -6.8 mmHg), indicating that the findings are robust to potential publication bias.

### BP reduction

Figure 1 shows the change in SBP from baseline to endpoint by treatment groups across all included trials. Overall, in the triple and dual combination groups, SBP reduction from baseline was 26.9 mmHg and 21.7 mmHg, respectively (difference: 5.4 mmHg; 95% CI: 4.7–6.2). In trials with an AHTD-free baseline (baseline mean SBP: 166 mmHg), SBP reduction was 34.3 mmHg and 29.5 mmHg in the triple and dual combination groups, respectively (difference: 5.2 mmHg; 95% CI: 4.2–6.1; p<0.0001). In trials with uncontrolled BP on dual combinations (baseline mean BP: 149 mmHg), SBP reduction was 14.7 mmHg and 8.6 mmHg in the triple and dual combination groups, respectively (difference: 6.1 mmHg; 95% CI: 4.4–7.8; p<0.0001). In the triple run-in trial (baseline mean BP: 133/81 mmHg), SBP reduction was 5.5 mmHg and 0.1 mmHg in the triple and dual combination groups, respectively (difference: 5.4 mmHg; 95% CI: 4.0–7.0; p<0.0001).

**Fig. 1.**
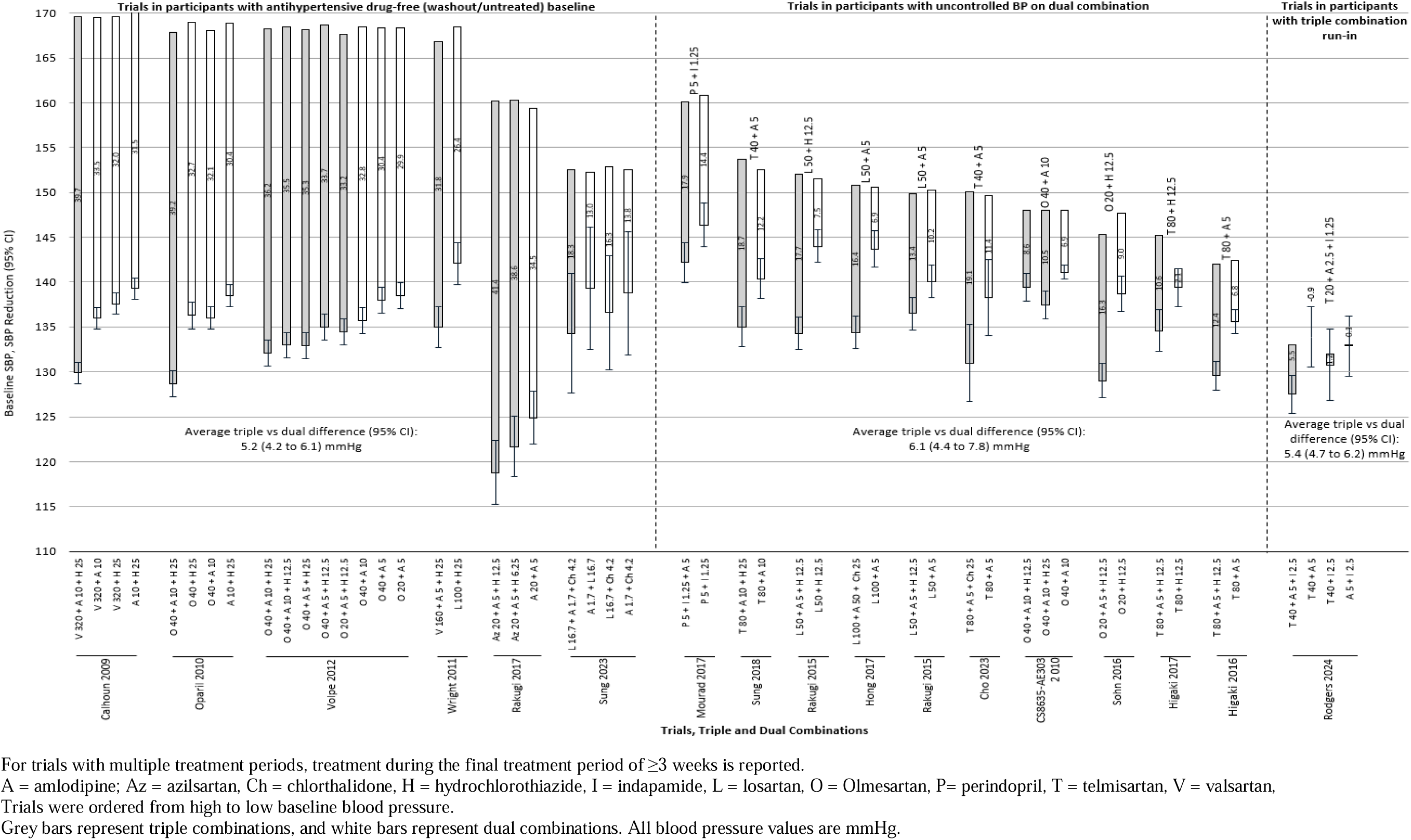
Baseline systolic blood pressure and corresponding systolic blood pressure reduction by trial, stratified by triple and dual combination types.

Heterogeneity in treatment effects was substantial. Subgroup analysis, based on the dose of the third drug in the triple combination, reduced heterogeneity in AHTD-free baseline trials but not in uncontrolled-on-dual trials. However, heterogeneity estimates were based on a small number of comparisons in each subgroup.

In the triple combination groups, compared to dual combination groups, the additional SBP reductions achieved with the third drug at standard doses of <0.5, 0.5, 1, and ≥2 were 4.0 mmHg (95% CI: 1.1–6.9), 3.6 mmHg (95% CI: 2.3–5.0), 6.3 mmHg (95% CI: 5.2–7.4), and 7.9 mmHg (95% CI: 6.6–9.1), respectively (Figure 2). These effects were similar across both the AHTD-free baseline and uncontrolled-on-dual trials **(Table 1)**. Among those on maximal dose dual combination, adding a third drug (forced up-titration) at standard dose, irrespective of BP, resulted in an additional SBP reduction of 7.1 mmHg compared to continuing the same regimen (95% CI:4.6–9.6; p<0.0001), while addition of a third AHTD at standard dose to maximum + standard, compared to continuing the same dual combination resulted in an additional SBP reduction of 7.2 mmHg (95% CI: 5.6–9.5; p<0.0001).[13] Among participants with uncontrolled BP despite treatment with dual combination therapy, adding a third drug consistently produced greater SBP reductions than further dose intensification of the existing two drugs. In trials where both drugs in the dual combination were already at maximal or standard dose, adding a third agent at half or standard dose yielded a statistically significant additional reduction in SBP compared with continuing the same dual regimen or simply increasing doses of the two drugs. Similarly, in those on sub-maximal dual therapy, the addition of a third drug at half or standard dose was associated with larger SBP reductions than intensifying dual therapy alone **(Supplementary Table S7)**. [14–20]

**Fig. 2.**
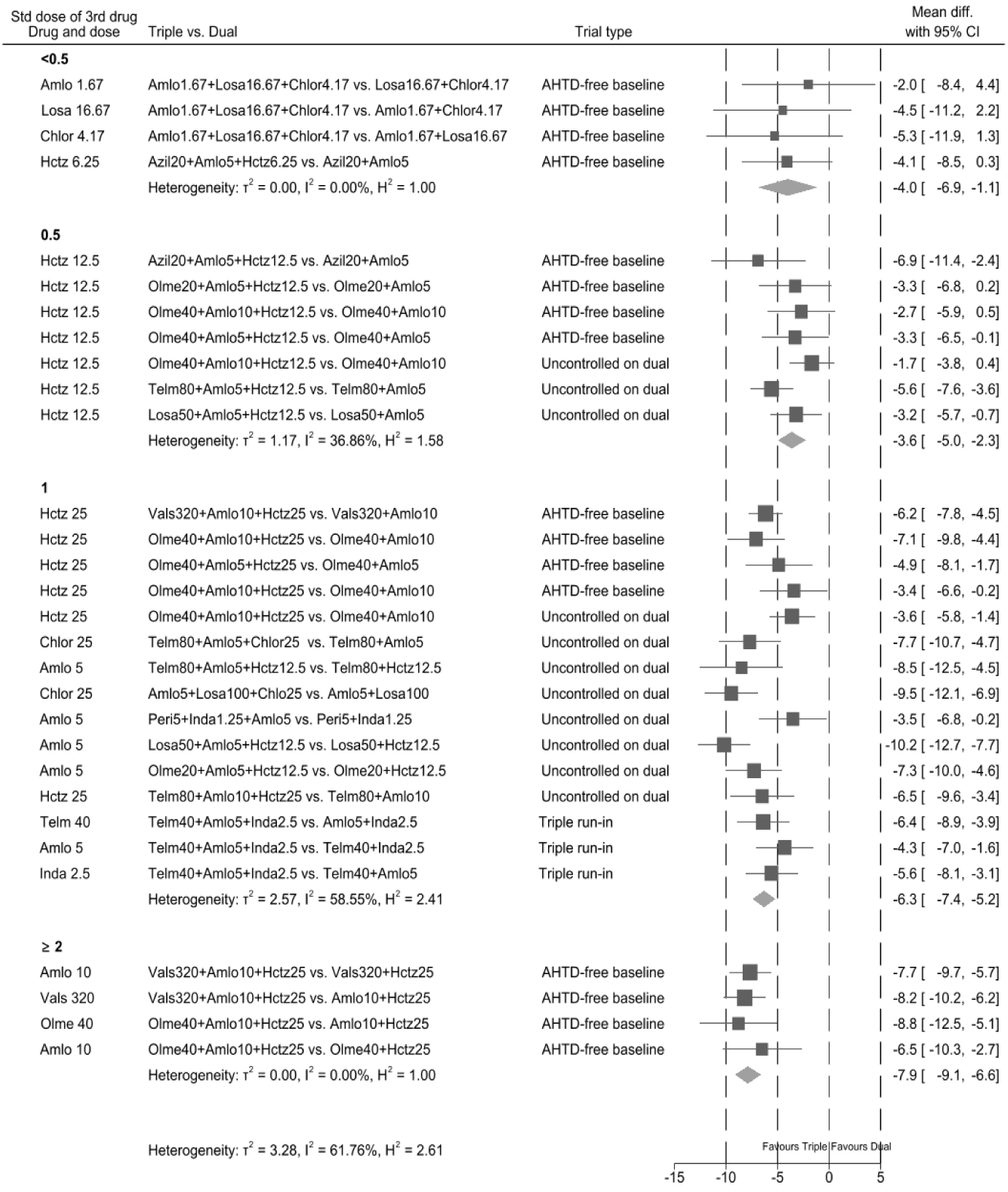
Difference in mean systolic blood pressure by standard dose of the third drug. Amlo- amlodipine, Azil- azilsartan, Losa- losartan, chlor- chlorthalidone, Hctz- hydrochlorothiazide, Olme- Olmesartan, Peri- Perindopril, Telm- telmisartan, Vals- Valsartan

**Table 1:** Blood pressure reduction outcome.

| Systolic blood pressure |  |  |  |  |  |  |  |  |  |
| --- | --- | --- | --- | --- | --- | --- | --- | --- | --- |
| Comparison type |  | Trials;<br>Comparisons | participants | Baseline<br>mean | Reduction<br>mean |  | Difference in the<br>reduction mean<br>(95% CIs) | P value | I<sup>2</sup> |
|  |  |  |  |  | Triple | Dual |  |  |  |
| AHTD-free baseline:<br>Overall |  | 6; 27 | 8329 | 166 | -34.3 | -29.5 | -5.2 (-6.1, -4.2) | <0.0001 | 36.9% |
| Standard<br>dose of the<br>third drug in<br>triple | <0.5 | 2; 4 | 351 | 154 | -23.4 | -19.4 | -4.0 (-6.9, -1.1) | 0.006 | 0% |
|  | 0.5 | 2; 4 | 637 | 166 | -36 | -32 | -3.7 (-5.4, -1.9) | <0.0001 | 0% |
|  | 1.0 | 3; 4 | 1943 | 169 | -37.6 | -32.2 | -5.8 (-7.0, -4.6) | <0.0001 | 0% |
|  | ≥2.0 | 2; 4 | 3130 | 169 | -39.5 | -31.7 | -7.9 (-9.1, -6.6) | <0.0001 | 0% |
| Uncontrolled-on-dual:<br>Overall |  | 11; 11 | 3644 | 149 | -14.7 | -8.6 | -6.1 (-7.8, -4.4) | <0.0001 | 76% |
| Standard<br>dose of the<br>third drug in<br>triple | <0.5 | NA | NA | NA | NA | NA | NA | NA | NA |
|  | 0.5 | 3; 3 | 1031 | 147 | -11.5 | -8 | -3.5 (-5.8, -1.2) | 0.003 | 70% |
|  | 1.0 | 8; 8 | 2613 | 150 | -15.9 | -8.8 | -7.1 (-8.9, -5.3) | <0.0001 | 69% |
|  | ≥2.0 | NA | NA | NA | NA | NA | NA | NA | NA |
| Triple run-in |  | 1; 3 | 1386 | 133 | -5.5 | -0.1 | -5.5 (-7.0, -4.0) | <0.0001 | 0% |

I<sup>2</sup> = heterogeneity, NA = Not Available All blood pressure values are in mmHg

Among participants with uncontrolled BP on one standard dose dual combination (both drugs at standard dose), doubling the dose of 1 drug in dual combination vs adding a third drug at the standard dose along with doubling the dose of one drug in the dual combination, resulted in an additional SBP reduction of 8.9 mmHg (95% CI: 6.9–10.8; p<0.0001).[21, 22]

Among participants with uncontrolled BP on standard dose dual combination, adding a third drug at the standard dose along with doubling the dose of both the drugs in the dual combination compared to doubling the dose of both the drugs in dual combination, resulted in an additional SBP reduction of 6.5 mmHg (95% CI: 3.4 to 9.6; p = <0.0001) mmHg [23]**(Supplementary Table S7)**.

From the one trial of triple combination run-in, after 6 weeks of randomized treatment with continued triple versus dual combinations, doubling the dose of all drugs in triple and dual combinations resulted in an additional SBP reduction difference of 0.8 mmHg (95% CI: -1.9 to 3.5, p = <0.56) [24]. Meta-regression analysis for the association between baseline SBP and SBP reduction suggested that every 10 mmHg increase in baseline SBP was associated with an additional 0.4 mmHg reduction in SBP for triples vs duals (R² = 1.2%). Overall, in the triple and dual combination groups, DBP reduced by 16.6 mmHg and 13.7 mmHg, respectively (difference: 3.2 mmHg; 95% CI: 2.6–3.8; p<0.0001). Subgroup analysis for DBP reported similar observations as that of SBP (Supplementary Table S6).

### Blood pressure control

Overall, BP control rates were 59% in the triple combination versus 46% in the dual combination group [risk ratio (RR 1.3 (95% CI: 1.27–1.40)), p<0.0001, I² = 54%]. The increase in BP control rates according to the dose of the third AHTD in the triple combination were [RR- 0.65 (95% CI: 0.47-0.83), RR- 0.68 (95% CI: 0.62-0.73) and RR- 0.70 (95% CI: 0.68-0.73)] for half, standard and 2+ standard doses, respectively.

BP control was higher in the triple vs dual groups in AHTD-free baseline trials (62% vs 49%), uncontrolled-on-dual trials (50% vs 35%) and the single triple run-in trial (74% vs 58%) (Table 2). While the absolute control rates differed across each set of trials, the risk differences were consistent with a 13-16% absolute improvement in BP control conferred by triple therapy, i.e. about 1 in 6 extra reaching BP control.

**Table 2:** Blood pressure control outcome.

| Comparison type |  | Trials;<br>Comparisons | Participants | % of participants with BP control<br><140/90mmHg |  | Relative risk<br>(95% CIs) | P value | I<sup>2</sup> (%) |
| --- | --- | --- | --- | --- | --- | --- | --- | --- |
|  |  |  |  | Triple (%) | Dual (%) |  |  |  |
| AHTD-free baseline<br>(Overall) |  | 6; 27 |  | 62 | 49 | 1.3 (1.3-1.4) | <0.0001 | 49 |
|  | <0.5 | 2; 4 | 384 | 74 | 57 | 1.1 (1.0-1.2) | <0.25 | 0 |
|  | 0.5 | 2; 4 | 2150 | 70 | 62 | 1.1 (1.1-1.2) | <0.0001 | 0 |
|  | 1.0 | 3; 4 | 3711 | 71 | 56 | 1.3 (1.2-1.4) | <0.0001 | 57 |
|  | ≥2.0 | 2; 4 | 3512 | 70 | 47 | 1.5 (1.3-1.7) | <0.0001 | 74 |
| Uncontrolled on dual<br>(Overall) |  | 11; 11 | 3621 | 50 | 35 | 1.5 (1.4-1.6) | <0.0001 | 65 |
| Standard<br>dose of<br>the third<br>drug in<br>triple | <0.5 | NA | NA | NA | NA | NA | NA | NA |
|  | 0.5 | 3; 3 | 1171 | 46 | 38 | 1.2 (1.0-1.4) | <0.012 | 21 |
|  | 1.0 | 8; 8 | 2719 | 52 | 31 | 1.7 (1.5-1.8) | <0.0001 | 11 |
|  | ≥2.0 | NA | NA | NA | NA | NA | NA | NA |
| Triple run-in |  | 1; 3 | 1384 | 74 | 58 | 1.3 (1.3-1.4) | <0.0001 | 27 |

I<sup>2</sup> = heterogeneity. BP control = systolic blood pressure <140 mmHg and diastolic blood pressure <90 mmHg

### Safety

Overall, from 15 trials (39 comparisons) reporting WDAE data, 4% and 3% of participants in the triple and dual combination groups, respectively, experienced WDAEs (RR=1.5 [95% CI 1.2–1.8], p=0.002). Further, 15 trials reported data on AAEs and TRAEs, and the incidence was higher in the triple combination group than the dual combination group for both AE [57% vs 29%; RR=1.8 (1.5–2.1)] and TRAE [24% vs 12%; RR=1.8(1.6–2.2)] **(Table 3)**.

**Table 3:**
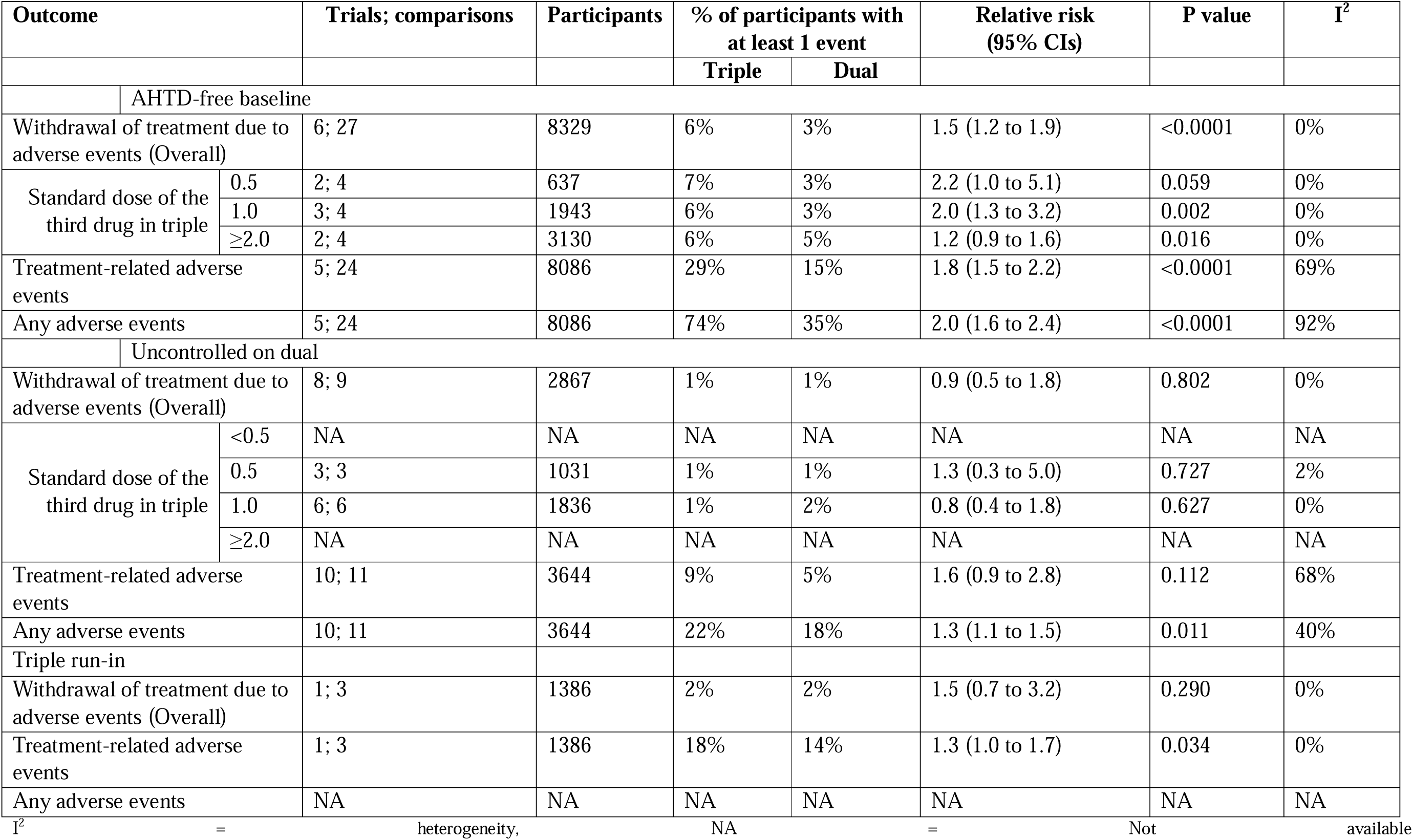
Safety outcomes.

In most of the included trials, adequate data on the specific types or severity of adverse events were limited. The higher incidence of AAEs and TRAEs in the triple therapy groups likely reflects expected class-related effects, such as peripheral oedema with calcium-channel blockers, symptoms related to greater blood pressure reduction (e.g., dizziness), and electrolyte changes associated with diuretics.

In trials with an AHTD-free baseline, the triple combination group showed a higher incidence of WDAEs, TRAEs, and AAEs compared to the dual combination group. In contrast, in uncontrolled-on-dual trials, no significant difference was observed between the dual and triple combination groups for any of the safety outcomes, except for AAE, while in a trial with triple run-in, no significant difference was observed between the dual and triple combination groups for any of the safety outcomes, except for TRAE **(Table 3)**.

### Certainty of evidence

The GRADE of evidence was categorized as “high” for both SBP and DBP outcomes, despite the high level of heterogeneity **(Supplementary Table S8**). The GRADE of evidence was categorized as “moderate” for the BP control outcome **(Supplementary Table S8)**. The GRADE of evidence was categorized as “low” for AE and TRAE and “very low” for WDAE (**Supplementary Table S8)**.

## Discussion

### Summary of key findings

This systematic review of double-blind randomized trials demonstrated that triple combinations of AHTDs conferred clinically relevant improvements in BP reduction and control compared to dual combinations. These effects were consistent in patients who were untreated and those uncontrolled on dual therapy. There was an increase in withdrawals in treatment due to adverse events in triple compared to dual combination groups in AHTD-free baseline trials, but not in those uncontrolled on dual combination therapy. Among participants with uncontrolled BP, addition of a third drug at half and one standard dose to a sub-maximal dual combination was found to be two times more efficacious when compared to addition of a third drug at half and one standard dose to a maximum dual dose combination.

Our analysis also demonstrates that adding a third AHTD to dual combination therapy results in greater SBP reductions than escalating doses of the existing drugs. This finding underscores the efficacy of expanding pharmacological diversity by targeting multiple mechanisms of action, especially in patients with uncontrolled blood pressure on standard-dose dual therapy.

The higher incidence of AAEs and TRAEs with triple therapy likely reflects predictable, class-related effects (e.g., peripheral oedema, dizziness, electrolyte disturbances). Although detailed adverse-event profiles were not consistently reported, these events seldom led to discontinuation. Nonetheless, this increased frequency of minor symptoms may influence tolerability and adherence in routine practice and should be considered when selecting treatment strategies.

Substantial between-study heterogeneity was observed for the primary and other outcomes. Baseline BP and treatment status, and the standard dose of the third drug, partly explained this variability. Other trial-level characteristics that varied across trials and that may have contributed to heterogeneity include: (i) differences in drug classes used within triple and dual combinations (e.g., variation in specific ARBs, CCBs, or diuretics and their pharmacodynamic profiles); (ii) differences in treatment duration ranging from 3 to 12 weeks; (iii) variability in participant characteristics such as age distribution, racial/ethnic composition, and prevalence of comorbidities such as diabetes.

### Comparison with evidence from previous research

Compared to our previous systematic review on this topic, this review included three additional trials involving 2,000 more participants and reported similar overall findings. Additionally, it evaluated the effects of up-titration of dual and triple combinations. Pragmatic randomized open-label trials [25, 26] reported the superiority of initial/early treatment with a low-dose triple SPC-based treatment protocol compared to usual care. A recent systematic review assessing the efficacy and safety of low-dose triple combination therapy in comparison to standard care, monotherapy, and placebo also concluded that triple pill therapy provided superior blood pressure control without a difference in the incidence of adverse events, aligning with our findings.[27]. Together with our results, this growing evidence base highlights the value of targeting multiple complementary mechanisms of action at lower individual doses and supports wider adoption of triple-combination strategies earlier in the treatment pathway.

### Implications

The proportion of patients receiving adequate hypertension care is suboptimal in the United States and many other countries. [28] Our review reinforces the recommendation that triple combination therapy should be used earlier, before dose intensification of dual therapy. Given the growing body of evidence on triple combination therapy, including RCTs on single-pill triple combinations, the global adoption and uptake of triple combination therapy for hypertension are likely to increase. This is crucial to address the rising prevalence of hypertension and improve very low BP control rates worldwide. Several newer single-pill triple combinations are currently under development. [24,29]

Current major hypertension guidelines, including the 2023 ESH and 2025 AHA/ACC guidelines, [4,5] recommend initiating treatment with dual combination therapy for most patients and subsequently intensification to triple therapy when BP remains above target. However, several guidelines still advise maximising the doses of dual therapy before introducing a third drug. Our findings directly challenge this paradigm. [30,31] Across trials, adding a third drug, even when patients were on standard-dose or submaximal dose dual therapy, produced substantially greater BP reductions than further dose intensification of the existing two drugs. These results support an earlier transition to triple therapy, consistent with the evolving approach favouring multi-mechanism treatment at lower individual drug doses to improve BP control.

### Strengths and limitations

The strengths of this systematic review include its inclusion of a larger number of trials and participants compared to previous reviews, providing up-to-date evidence on the effects of triple combinations of AHTDs. The review also assessed dose-doubling effects in forced up-titration trials, offering novel insights into the impact of dose intensification. Heterogeneity in treatment effects was partly explained by baseline BP and treatment status, and the standard dose of drugs. Residual heterogeneity was high, and other factors that contributed to heterogeneity could not be formally examined because subgroup sample sizes were limited. This reduces the generalisability and precision of the findings, and it is reflected in the GRADE assessments, where evidence certainty was downgraded. Further, a detailed assessment of specific adverse event types was not possible as most trials did not report AAEs or TRAEs types or their severity in sufficient detail.

### Future direction

Future research should focus on evaluating long-term outcomes, including the impact on cardiovascular events, to provide more definitive guidance for clinical practice. Additionally, implementation research is essential to assess the feasibility, acceptability, and sustainability of integrating triple combination therapy into routine practice. This involves understanding barriers to uptake, designing effective implementation strategies, and addressing therapeutic inertia among healthcare providers and patients. Cost-effectiveness analyses are equally crucial, particularly in low-and middle-income countries where accessibility and affordability pose significant challenges. Collaborative efforts involving clinicians, researchers, and policymakers will be vital to ensure that emerging evidence translates effectively into real-world settings, ultimately optimising hypertension control on a global scale.

## Conclusion

Treatment with triple combinations of AHTDs is more efficacious compared to dual combinations and well-tolerated. There is an inconsistency across clinical guidelines recommending the use of triple therapy for hypertension, with some recommending it only after maximal dose dual therapy fails and others suggesting earlier use. This review advocates for earlier initiation of triple combination therapy, prior to maximal dose dual therapy, to achieve lower BP targets of <130/80mmHg in most patients.

## Data Availability

All data produced in the present study are available upon reasonable request to the authors

## Contributions

AS and RD contributed to the conception, data acquisition, data interpretation, and drafting of the manuscript. GS was involved in data acquisition, data interpretation, and drafting the manuscript. RP, AM were responsible for data analysis and interpretation. AR contributed to the conception and data interpretation. All authors critically reviewed the manuscript and approved it for publication.

## Funding

Program grant from the National Health and Medical Research Council, Australia: GNT1149987, and partly funded by DBT/Wellcome Trust India Alliance Fellowship [Fellowship number IA/CPHI/20/1/505281].

## Role of funders

Funders had no role in designing, conducting, analysing, or reporting of this work.

## Statements and Declarations

Competing interests: AR is employed by The George Institute for Global Health (TGI) and Imperial College London. TGI has submitted patent applications for low-dose combination products for hypertension, and AR is listed as an inventor. George Medicines Pty Ltd (GM) is a subsidiary of TGI, holds a license for these patents and has received investment to develop these combination therapies. AR is seconded part-time to GM. None of the authors have financial interest in these patents or in GM.

## Data Availability

The data sets that were generated during the analysis will be available from the corresponding author on reasonable request.

