## Supplementary file for "Efficacy and safety of triple versus dual combinations of antihypertensive drugs: A systematic review and meta-analysis of double-blind randomized clinical trials"

**Table S1: PubMed search**

| **Search number** | **Query** | **Results** |
| --- | --- | --- |
| 24 | #3 AND #6 AND #17 AND #21, Humans, from 2019/1/1 - 2024/10/31 | 339 |
| 23 | #3 AND #6 AND #17 AND #21, Humans | 2,731 |
| 22 | #3 AND #6 AND #17 AND #21 | 2,856 |
| 21 | #18 or #19 or #20 | 1,572,330 |
| 20 | random*[Title/Abstract] | 1,567,307 |
| 19 | exp Random Allocation/ | 2,740 |
| 18 | exp Randomized Controlled Trial/ | 8,631 |
| 17 | #7 OR #8 OR #9 OR #10 OR #11 OR #12 OR #13 OR #14 OR #15 OR #16 | 828,555 |
| 16 | polypharmacy/ | 15,460 |
| 15 | Drug Therapy, Combination/ | 552,468 |
| 14 | Single pill | 3,318 |
| 13 | Polycap | 32 |
| 12 | Polypill | 749 |
| 11 | Fixed combination | 46,547 |
| 10 | Fixed dose* | 33,145 |
| 9 | Multidrug | 103,127 |
| 8 | Quadruple | 9,328 |
| 7 | Triple | 117,630 |
| 6 | #4 OR #5 | 684,213 |
| 5 | hypertensi* | 662,152 |
| 4 | exp Vascular Diseases/ | 30,921 |
| 3 | #1 OR #2 | 117,871 |
| 2 | antihypertensive* | 107,963 |
| 1 | exp Antihypertensive Agents/ | 12,279 |

**Table S2: Cochrane Central Register of Controlled Trials**

| Search number | Query | Results |
| --- | --- | --- |
| 1 | exp Antihypertensive Agents/ | 35967 |
| 2 | antihypertensive*.tw. | 14430 |
| 3 | 1 or 2 | 43597 |
| 4 | exp Vascular Diseases/ | 116962 |
| 5 | hypertensi*.tw. | 68645 |
| 6 | 4 or 5 | 160768 |
| 7 | triple.tw. | 15526 |
| 8 | Quadruple.tw. | 1804 |
| 9 | multidrug.tw. | 1964 |
| 10 | Fixed dose*.tw. | 8461 |
| 11 | Fixed combination.tw. | 1901 |
| 12 | Polypill.tw. | 170 |
| 13 | Polycap.tw. | 22 |
| 14 | Single pill.tw. | 349 |
| 15 | Drug Therapy, Combination/ | 37035 |
| 16 | polypharmacy/ | 410 |
| 17 | or/7-16 | 63193 |
| 18 | exp Randomized Controlled Trial/ | 37 |
| 19 | exp Random Allocation/ | 26032 |
| 20 | random*.tw. | 1293163 |
| 21 | or/18-20 | 1299608 |
| 22 | 3 and 6 and 17 and 21 | 2909 |
| 23 | limit 22 to yr="2019 -Current" | 263 |

**Table S3: Standard dose of antihypertensive drugs**

| Antihypertensive drug | WHO DDD | BNF | Martindale | MIMS online | STANDARD DOSE |
| --- | --- | --- | --- | --- | --- |
| Amlodipine | 5 mg | 5 - 10 mg | 5 - 10 mg | 2.5 - 10 mg | 5 mg |
| Azilsartan | - | 40 mg | 40 mg | - | 40 mg |
| Chlorthalidone | 25 mg | - | 25 mg | - | 25 mg |
| Hydrochlorothiazide | 25 mg | - | 25-50 mg | - | 25 mg |
| Indapamide | 2.5 mg | 2.5 mg | 1.25-2.5 mg | 1.5 mg | 2.5 mg |
| Losartan | 50 mg | 50 mg | 50 mg | 50 mg | 50 mg |
| Perindopril | 5 mg | 5 mg | 5-10 mg | - | 5 mg |
| Olmesartan | 20 mg | 10-20 mg | 20 mg | 20 mg | 20 mg |
| Telmisartan | 40 mg | 40 mg | 20-80 mg | 40 mg | 40 mg |
| Valsartan | 80 mg | 80 mg | 80-160 mg | 80 mg | 80 mg |

BNF=British National Formulary; DDD=Defined daily dose; Monthly Index of Medical Specialties, Australia; WHO=World Health Organization.

Standard dose is defined as the most reported usual maintenance dose given in the British National Formulary Martindale and Monthly Index of Medical Specialties, Australia. In case of no consensus from these sources, the World Health Organization’s defined daily dose was used as a tiebreaker.

**Table S4: Summary of characteristics of included trials**

| **Author/Study, Year** | **Key eligibility** | **Sample size** | **Concomitant conditions** | **Washout/ placebo run in weeks** | **BP measure** | **SBP, mmHg** | **DBP, mmHg** | **Regimen** | **Treatment duration, weeks** |
| --- | --- | --- | --- | --- | --- | --- | --- | --- | --- |
| **AHTD -free baseline trials** | | | | | | | | | |
| Calhoun, 2009 | SBP 145-199 & DBP 100-119 after 1-4 weeks on placebo | 2271 | NR | 1 to 4 | Seated, trough | 170 | 106 | Forced uptitration | 4 |
| Oparil, 2010 | SBP ≥140 or DBP ≥100 (or ≥160 or ≥90 off- treatment) | 2492 | DM 16% CKD 4%  CVD 9% | 3 | Seated, trough | 169 | 101 | Forced uptitration | 8 |
| Wright, 2011 | SBP 160-199 after 1-2 weeks of washout | 488 | DM 20% | 1 to 2 | Seated, NR | 168 | 98 | Forced uptitration | 3 |
| Volpe, 2012 | SBP 160-199 & DBP 100-114 after 2 weeks washout | 2690 | DM 15% CKD 2%  CVD 29% Obese 51% | 3 | Seated, trough | 168 | 104 | Forced uptitration | 8 |
| Rakugi, 2017 | SBP 150-179, DBP 95-109 after 4 weeks on placebo | 209 | NR | 4 | Seated, trough | 161 | 100 | Forced uptitration | 8 |
| Sung, 2023 | SBP ≥140-<180, DBP <110 after 4 weeks on placebo | 245 | DM 61% | 4 | Seated, trough | 153 | 92 | Fixed | 8 |
| **Uncontrolled on dual trials** | | | | | | | | | |
| CS8635-A-E303, 2010 | SBP 140-200 & DBP 90-115 after 4 weeks on Olme 40 + Amlo 10 | 808 | NR | No | Seated, trough | 148 | 94 | Fixed | 8 |
| Rakugi, 2015-1 | DBP 90-109 & SBP 140-199 after 8 weeks on Losa 50 + Hctz 12.5 | 286 | DM 16% | No | Seated, trough | 152 | 96 | Fixed | 8 |
| Rakugi, 2015-2 | DBP 90-109 & SBP 140-199 after 8 weeks on Losa 50 + Amlo 5 | 327 | DM 13% | No | Seated, trough | 150 | 96 | Fixed | 8 |
| Higaki, 2016 | DBP 90-114, SBP ≤200, after 6 weeks on Telm 80 + Amlo 5 | 309 | NR | No | Seated, trough | 142 | 96 | Fixed | 8 |
| Sohn, 2016 | SBP >140/90 (>130/80 for DM/CKD) after 4 weeks on Olme 20 + Hctz 12.5 | 341 | DM 20%  CKD 11% | No | Seated, NR | 147 | 94 | Fixed | 8 |
| Higaki, 2017 | DBP 90-114, SBP ≤200, after 6 weeks on Telm 80 + Hctz 12.5 | 132 | NR | No | Seated, trough | 143 | 97 | Fixed | 8 |
| Hong, 2017 | SBP 140-199 after 4 weeks on Amlo 5 + Losa 50 | 340 | DM 17% | No | Seated, NR | 151 | 92 | Forced uptitration | 6 |
| Mourad, 2017 | BP ≥150/95 after 1 month on peri 5 + Inda 1.25 | 454 | NR | No | Seated, trough | 160 | 101 | Fixed | 4 |
| Sung, 2018 | SBP 140-199 or (130-199 for DM/CKD) after 4 weeks on Telm 40 + Amlo 5 | 310 | DM 26% CKD 15% | No | Seated, NR | 153 | 89 | Forced uptitration | 8 |
| Cho, 2023 | SBP140-≤200, T2DM or CKD-SBP130-≤200 after 4 weUeks on Telm 40 + Amlo 5 | 374 | DM-24% CKD- 14% Dyslipidemia-49% | No | Seated, NR | 150 | 89 | Forced uptitration | 8 |
| **Triple run-in** | | | | | | | | | |
| Rodgers, 2024 | SBP 140-179 (no drugs), SBP 130-170 (1 drug), SBP 120-160 (2 drugs), SBP 110-150 (3 drugs) | 1385 | DM-22% Dyslipidemia-46% | No | Seated, trough | 142 | 85 | Forced uptitration | 12 |

A = amlodipine; AHTD = Anti-hypertensive Drug Az = azilsartan, Ch = chlorthalidone, CKD = Chronic Kidney Disease, CVD = Cardiovascular Disease, DBP = Diastolic Blood Pressure, H = hydrochlorothiazide, I = indapamide, L = losartan, O = Olmesartan, P= perindopril, SBP = Systolic Blood Pressure, T = telmisartan, T2(DM) = Type -2 (Diabetes Mellitus), V = valsartan

**Table S5: Risk of bias in included trials**

| Trial Year | RAND Seq Generation | Allocation concealment | Blinding of participants and personnel | Blinding of outcomes assessors | Incomplete outcome data | Selective reporting of outcomes | Other Bias |
| --- | --- | --- | --- | --- | --- | --- | --- |
| Calhoun et al. 2009 | L | L | L | L | L | L | L |
| CS8635-AE303 2010 | L | L | L | L | L | L | L |
| Higaki et al. 2016 | L | L | L | L | L | L | U |
| Higaki et al. 2017 | L | L | L | L | L | L | U |
| Hong et al. 2017 | L | L | L | L | L | L | L |
| Mourad et al. 2017 | L | L | L | L | L | L | L |
| Oparil et al. 2010 | L | L | L | L | L | L | L |
| Rakugi et al. 2017 | L | L | L | L | L | L | U |
| Rakugi et al.a 2015 | L | L | L | L | L | L | U |
| Rakugi et al.b 2015 | L | L | L | L | L | L | U |
| Sohn et al. 2016 | L | L | L | L | L | L | L |
| Sungv et al. 2018 | L | L | L | L | L | L | L |
| Volpe et al. 2012 | L | L | L | L | L | L | L |
| Wright et al. 2011 | L | L | L | L | L | L | L |
| Cho et al. 2023 | L | L | L | L | L | L | L |
| Sung et al. 2023 | L | L | L | L | L | L | L |
| Rodgers et al. 2024 | *L* | L | L | L | L | L | L |

L = low-risk of bias; *L* = inadequate information reported to make a judgement, but most probably low-risk of bias; U = unclear risk of bias

**Table S6: Diastolic blood pressure reduction outcomes**

| **Diastolic blood pressure** | | | | | | | | |
| --- | --- | --- | --- | --- | --- | --- | --- | --- |
| **Comparison type** | | **Trials;**  **Comparisons**  **(participants)** | **Baseline mean** | **Reduction**  **mean** | | **Difference in reduction mean**  **(95% CIs)** | **P value** | **I^2^** |
|  | |  |  | Triple | Dual |  |  |  |
| AHTD-free baseline (Overall) | | 6; 27 (8329) | 101 | -20.1 | -17.4 | -3.0 (-3.7, -2.3) | <0.0001 | 55% |
| Standard dose of third drug in triple | <0.5 | 2; 4 (351) | 94 | -8.6 | -6.4 | -2.1 (-4.3, 0.02) | 0.048 | 0% |
|  | 0.5 | 2; 4 (637) | 91 | -19.2 | -16.5 | -2.7 (-3.8, -1.6) | <0.0001 | 0% |
|  | 1.0 | 3; 4 (1943) | 104 | -23.3 | -20.4 | -3.2 (-3.8, -2.5) | <0.0001 | 0% |
|  | ≥2.0 | 2; 4 (3130) | 104 | -22.5 | -19.8 | -5.2 (-5.9, -4.5) | <0.0001 | 0% |
| Uncontrolled on dual  (Overall) | | 11; 11 (3644) | 94 | -12.3 | -8.7 | -3.5 (-4.7, -2.4) | <0.0001 | 81% |
| Standard dose of third drug in triple | 0.5 | 3; 3 (1031) | 95 | -8.3 | -6.2 | -2.1 (-4.0, -0.2) | 0.033 | 81% |
|  | 1.0 | 8; 8 (2613) | 94 | -13.8 | -9.6 | -4.1 (-5.4, -2.8) | <0.0001 | 78% |
|  | ≥2.0 | NA | NA | NA | NA | NA | NA | NA |
| Triple run-in | | 1; 3 (1386) | 81 | -3.4 | -0.6 | -4.0 (-5.0, -2.9) | <0.0001 | 0% |

I^2^ = heterogeneity, NA = Not Available

All blood pressure values are in mmHg

**Table S7: Interventions, comparators and titrations in the included trials**

| **Trials** | **Pre-randomization** | **Randomization** | | **Post randomization treatment** | **Inclusion/Exclusion in titration analysis**  **Results** |
| --- | --- | --- | --- | --- | --- |
|  | **Run- in** | **Period 1** | **Period 2** | **Period 3** |  |
| **AHTD -free baseline trials** | | | | | |
| Calhoun 2009 | 3 weeks  placebo | 1 week (2 vs 2)  A5 + H12.5  V160 + H 12.5  A5 + V160  V160 + H 12.5 | 1 week (dual vs triple)  Same dual  Same dual  Same dual  Same + A5 | 3 weeks (2 vs 3)  A10 + H25  V320 + H25  A10 + V320  A10 + V320 + H25 | Excluded.  Variable run-in duration and <3 weeks duration of period 1 and period 2. |
| Oparil  2010 | 0-3 weeks washout | 4 weeks (2 vs 2)  O40 + A10*  O40 + H25*  A10 + H25*  O40 + A10* | 8 weeks (2 vs 3)  Same dual  Same dual  Same dual  Same + H25 |  | Included forced up-titration from period 1 to period 2.  In those on dual combination therapy, continuing the same vs addition of a 3^rd^ drug at std dose to one of the dual combinations resulted in SBP reduction of 7.6 (6.0-9.1) mmHg (p = <0.0001).  Among those on max + std dose dual, addition of a 3^rd^ drug at std dose resulted in additional SBP reduction of 7.2 mmHg (95% CI: 5.6 to 9.5; p = <0.0001  Among those on max dual, addition of a 3rd drug at std dose resulted in additional SBP reduction of 7.1 mmHg (95% CI:4.6 to 9.6; p = <0.0001) |
| Wright 2011 | 1 week  washout | 3 weeks (2 vs 1)  V160 + A5  L100 | 3 weeks (3 vs 2)  Same + H25  Same + H25 |  | Included forced up titration from period 1 to period 2.  In those on dual and monotherapy, addition (forced up titration) of 3^rd^ drug and 2^nd^ drug at std dose respectively resulted in additional SBP reduction of 5.5 (2.3-8.7) mmHg (p = <0.001). |
| Volpe  2012 | 2 weeks  washout | 2 weeks (2 vs 2)  Placebo  O20 + A5  O40 + A5  O40 + A10 | 8 weeks (2 vs 3)  Same  Same  Same  O20 + A5 + H12.5  O40 + A5 + H12.5  O40 + A5 + H25  O40 +A10 + H12.5  O40 + A10 + H25 |  | Excluded.  <3 weeks duration of run-in and period 1. |
| Rakugi 17 | 4 weeks  placebo | 2 weeks (2 vs 2)  Az20 + A5 | 8 weeks (2 vs 3)  Same  Same + H 12.5  Same + H 6.25 |  | Excluded.  <3 weeks duration of period 1. |
| Sung 2023 | 4 weeks  placebo | 8 weeks (2 vs 3)  Amlo 1.67+Losa 16.67 Losa 16.67+Chlor 4.17 Amlo 1.67+Chlor 4.17  Amlo 1.67+Losa 16.67+Chlor 4.17 |  |  | Excluded.  No up-titration. |
| **Uncontrolled on dual trials** | | | | | |
| CS8635-A-E303 2010 | 4 weeks  O40 + A10 | 8 weeks (2 vs 2)  O40 + A10 O40 + A10 + H12.5 O40 + A10 + H25 |  |  | Included uptitration from run-in to period 1.  In those with uncontrolled BP on max + max dual, continuing same vs adding a 3rd drug at ½ std and std. dose resulted in additional SBP reduction of 1.7 (0.4-3.8) (p = <0.201), 3.6 (1.4-5.8) (p = < 0.007) mmHg |
| Hong  2017  Cho  2023  Sung  2018 | 4 weeks  A5 + L50  4 weeks  T40 + A5  4 weeks  T40 + A5 | 2 weeks (2 vs 3)  Same + Ch12.5  Same  2 weeks (2 vs 3)  Same + Ch12.5  Same  2 weeks (2 vs 3)  Same + H12.5  Same | 4 weeks (2 vs 3)  A5 + L100 + Ch25  A5+ L100  6 weeks (2 vs 3)  T80 + A5 + Ch25  T80 + A5  6 weeks (2 vs 3)  T80 + A10 + H25  T80 + A10 |  | Included up-titration from run-in to period 2.  In those uncontrolled on standard dose dual combination,   addition of a 3^rd^ drug at standard dose and doubling the dose of one drug in triple vs doubling the dose of one drug in dual  addition of a 3^rd^ drug at standard dose and doubling the dose of both drugs in triple vs doubling the dose of both drugs in dual  resulted in additional SBP reduction of 8.9 (6.9-10.8) mmHg, (p = <0.0001), 6.5 (3.4-9.6) (p = <0.0001)  Period 1 excluded because of <3 weeks duration. |
| Higaki  2017  Higaki  2016  Rakugi  2015-1  Sohn  2016  Mourad  2017  Rakugi 2015-2 | 6 weeks  T 80 + H12.5  6 weeks  T80 + A5  8 weeks  L50 + H 12.5  4 weeks  O20 + H12.5  4 weeks  P5 + I1.25  8 weeks  L50 + A5 | 8 weeks (2 vs 3)  Same + A5  Same  8 weeks (2 vs 3)  Same + H12.5  Same  8 weeks (2 vs 3)  Same + A5  Same  8 weeks (2 vs 3)  Same + A5  Same  4 weeks (2 vs 3)  Same + A5  Same  8 weeks (2 vs 3)  Same + H 12.5  Same |  |  | In those uncontrolled on sub-maximal dose dual combination, adding a third drug at half and one standard dose resulted in additional SBP reduction of 4.5 (2.2-6.9), 7.5 (4.7-10.2) mmHg (p- value for both 0.0001) |
| Triple run-in | | | | | |
| Rodgers  2024 | 4 weeks  T20 + A2.5 + I1.25 | 6 weeks (2 vs 3)  Same  A2.5 + I1.25  T20 + I1.25  T20 + A2.5 | 6 weeks (2 vs 3)  T40 + A5 + I2.5  A5 + I2.5  T40 + I2.5  T40 + A5 |  | Included. Forced uptitration from period 1 to period 2.  Doubling the dose of all drugs in triple vs both drugs in dual resulted in additional SBP reduction of -4.7, -3.9, -4.1, 3.6 mmHg (p = <0.0001). |
| **#** 2% patients were on placebo for 2 weeks before they received dual combination for 2 weeks.  A = amlodipine; Az = azilsartan, Ch = chlorthalidone, H = hydrochlorothiazide, I = indapamide, L = losartan, O = Olmesartan, P= perindopril, SBP= Systolic Blood Pressure, T = telmisartan, V = valsartan | | | | | |

| **Certainty assessment** | | | | | | | **№ of patients** | | **Effect** | | **Certainty** | **Importance** |
| --- | --- | --- | --- | --- | --- | --- | --- | --- | --- | --- | --- | --- |
| **No. of studies** | **Study design** | **Risk of bias** | **Inconsistency** | **Indirectness** | **Imprecision** | **Other considerations** | **Triple combinations** | **Dual combinations** | **Relative (95% CI)** | **Absolute (95% CI)** |  |  |
| **Reduction in SBP** | | | | | | | | | | | | |
| 17 | Randomised trials | Not serious | Serious* | Not serious | Not serious | Strong association dose response gradient | 5802 | 7557 | - | MD **5.4 mm Hg lower** (6.2 lower to 4.7 lower) | ⨁⨁⨁⨁ High | CRITICAL |
| **Reduction in DBP** | | | | | | | | | | | | |
| 17 | Randomised trials | Not serious | Serious* | Not serious | Not serious | Strong association | 5802 | 7557 | - | MD **3.2 mm Hg lower** (3.8 lower to 2.7 lower) | ⨁⨁⨁⨁ High | CRITICAL |
| **Proportion achieving BP Control** | | | | | | | | | | | | |
| 13 | Randomised trials | Not serious | Serious* | Not serious | Not serious | None | 2774/4020 (69.0%) | 3368/6354 (53.0%) | **RR 1.34** (1.27 to 1.41) | **180 more per 1,000** (from 143 more to 217 more) | ⨁⨁⨁◯ Moderate | CRITICAL |
| **Any adverse event** | | | | | | | | | | | | |
| 15 | Randomised trials | Not serious | Serious* | Not serious | Serious ^¥^ | None | 2957/5188 (57.0%) | 1897/6542 (29.0%) | **RR 1.8** (1.5 to 2.1) | **232 more per 1,000** (from 145 more to 319 more) | ⨁⨁◯◯ Low | IMPORTANT |
| **Treatment-related adverse events** | | | | | | | | | | | | |
| 13 | Randomised trials | Not serious | Serious* | Not serious | Serious ^¥^ | None | 1247/5197 (24.0%) | 821/6838 (12.0%) | **RR 1.8** (1.6 to 2.2) | **96 more per 1,000** (from 72 more to 144 more) | ⨁⨁◯◯ Low | IMPORTANT |
| **Withdrawal due to adverse events** | | | | | | | | | | | | |
| 15 | Randomised trials | Very serious | Serious* | Not serious | Serious ^¥^ | None | 190/4740 (4.0%) | 203/6770 (3.0%) | **RR 1.4** (1.2 to 1.8) | **12 more per 1,000** (from 6 more to 24 more) | ⨁◯◯◯ Very low | CRITICAL |
| *Inconsistency was marked as “serious” when a high level of heterogeneity was observed.  ^¥^Imprecision was marked as “serious” when a wide confidence interval (CI) was observed around the estimate of the effect.  **CI:** confidence interval; **MD:** mean difference; **RR:** risk ratio | | | | | | | | | | | | |

**Table S8: Certainty of evidence assessment using GRADEpro**

**Figure S1: PRISMA flow chart**

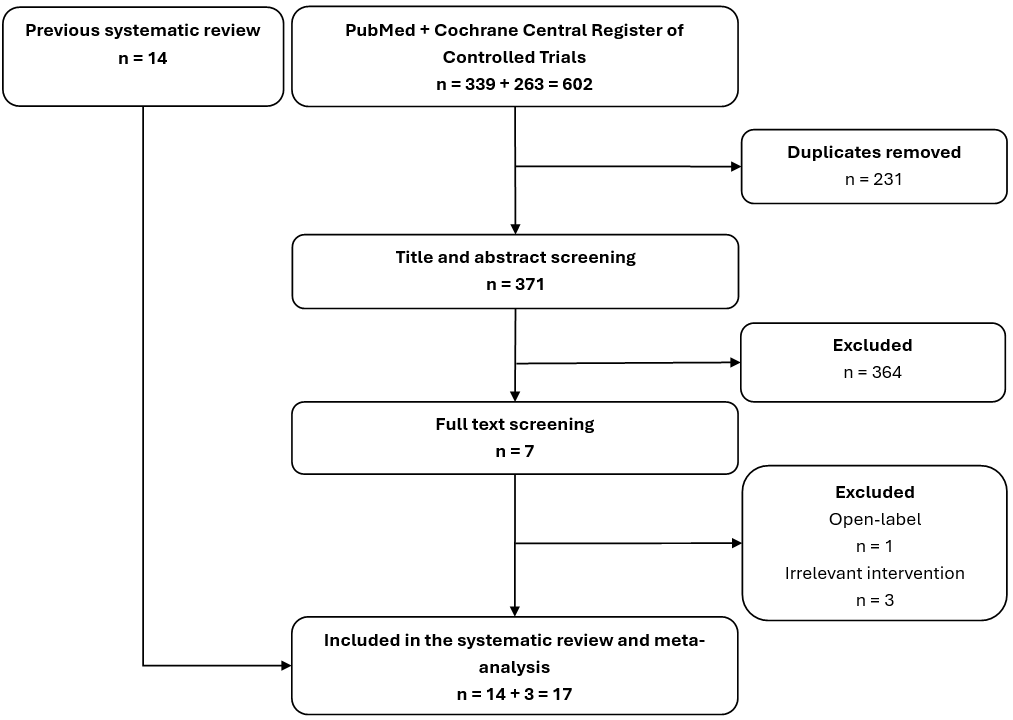

**SFigure2: Funnel plot for risk of bias assessment for primary outcome of systolic blood pressure reduction**

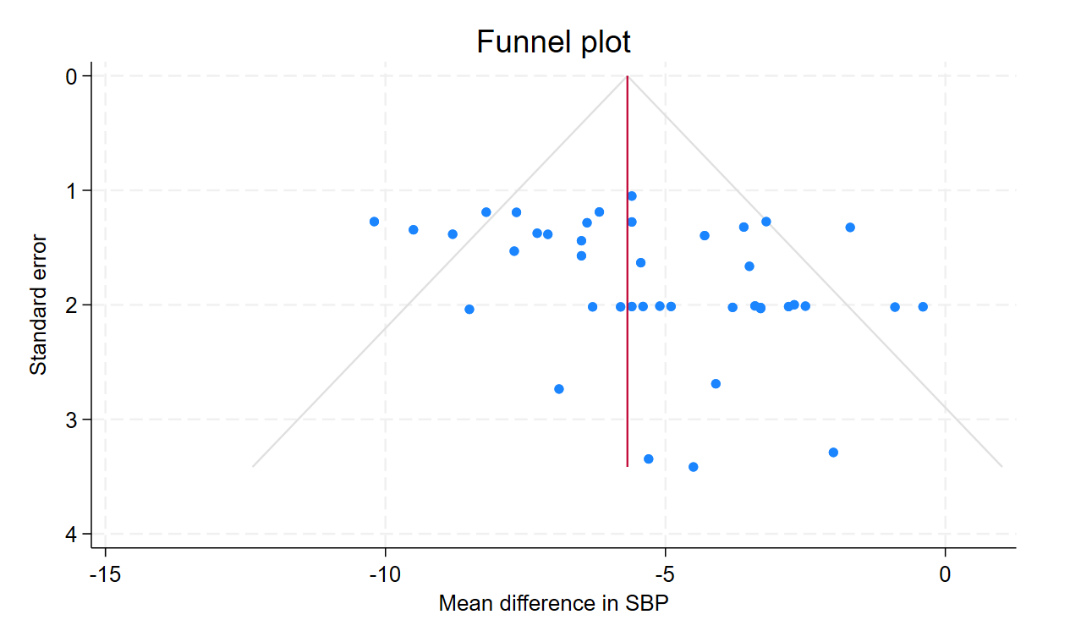
